# Mortality and Severity in COVID-19 Patients on ACEIs & ARBs - A Meta-Regression Analysis

**DOI:** 10.1101/2021.03.14.21253557

**Authors:** Romil Singh, Sawai Singh Rathore, Hira Khan, Abhishek Bhurwal, Mack Sheraton, Prithwish Ghosh, Sohini Anand, Janaki Makadia, FNU Ayesha, Kiran S. Mahapure, Ishita Mehra, Aysun Tekin, Rahul Kashyap, Vikas Bansal

**Affiliations:** Department of Anesthesiology and Critical Care Medicine, Mayo Clinic, Rochester, Minnesota, USA; Medical Student, Dr. Sampurnanand Medical College and Hospital, Jodhpur, Rajasthan, India; Department of Internal Medicine, Islamic International Medical College, Rawalpindi, Pakistan; Department of Gastroenterology and Hepatology, Rutgers Robert Wood Johnson School of Medicine, New Brunswick, NJ, USA; Department of Emergency Medicine, Trinity West Medical Center, Steubenville, Ohio, USA; Department of Gastroenterology and Hepatology, Mayo Clinic, Rochester, Minnesota, USA; Medical Graduate, Patliputra Medical College and Hospital, Dhanbad, Jharkhand, India; Medical Graduate, GMERS Medical College and Hospital, Gotri, Vadodara, Gujrat, India; Medical Graduate, Services Institute of Medical Sciences, Lahore, Pakistan; Department of Plastic Surgery, KAHER J. N. Medical College, Belgaum, Karnataka, India; Departments of Internal Medicine, North Alabama Medical Center, Florence, AL, USA; Department of Pulmonary and Critical Care Medicine, Mayo Clinic, Rochester, Minnesota, USA

**Keywords:** COVID-19, Angiotensin inhibitors, ACEI, ARB, Mortality, Severity, Meta-Analysis, Meta-Regression

## Abstract

**Purpose:** The primary objective of this review is to examine studies reporting association of mortality in COVID-19 patients with whether they were on Angiotensin-converting-enzyme inhibitors (ACEIs) and Angiotensin II receptor blockers (ARBs). A secondary objective is to similarly access associations with higher severity of the disease in COVID-19 patients.

**Materials and Methods:** We searched multiple COVID-19 databases (WHO, CDC, LIT-COVID) for randomized trials and longitudinal studies from all over the world reporting mortality and severity published before January 18^th^, 2021. Meta-analyses were performed using 53 studies for mortality outcome and 43 for the severity outcome. Mantel-Haenszel odds ratios were generated to describe overall effect size using random effect models. To account for between study results variations, multivariate meta-Regression was performed with preselected covariates using maximum likelihood method for both the mortality and severity models.

**Result:** Our findings showed that the use of ACEIs/ARBs did not significantly influence either mortality (OR=1.16 95% CI 0.94 to 1.44, p= 0.15, I^2^ = 93.2%) or severity (OR=1.18, 95% CI 0.94 to 1.48 p= 0.15, I^2^ = 91.1%) in comparison to not being on ACEIs/ARBs in COVID-19 positive patients. Multivariate meta-regression for the mortality model demonstrated that 36% of between study variations could be explained by differences in age, gender, and proportion of heart diseases in the study samples. Multivariate meta-regression for the severity model demonstrated that 8% of between study variations could be explained by differences in age, proportion of diabetes, heart disease and study country in the study samples.

**Conclusion:** We found no association of mortality or severity in COVID-19 patients taking ACEIs/ARBs.

## Introduction

SARS-CoV-2 originated in Wuhan, China, in December 2019 and has spread to every major country in the world and was subsequently declared a pandemic on March 11, 2020^1^. As of March 10^th^, 2021, there are 117,997,454 positive patients worldwide; and 2,619,379 of these patients were reported to be deceased because of SARS-CoV-2^2^. The case fatality rate of SARS-CoV-2 in the U.S. is 1.05 per 1000 population as per COVID-19 Dashboard by the Center for Systems Science and Engineering (CSSE) at Johns Hopkins University^2^. However, the role of different medications and comorbidities has been elicited in recent articles.

The SARS-CoV-2 disease varies from mild to fulminant in reference to several risk variables contributing to a poor prognosis^3-5^. While the virus significantly impacts the respiratory tract, other metabolic systems have been involved in numerous case studies and systematic reviews^6-14^. Thorough awareness of the risks, pathogenesis, and predisposing factors together with the important aspects in the diagnosis is of paramount importance in order to direct decision-making for acute care and mitigate mortality of COVID-19^15-19^.

Severe acute respiratory syndrome coronavirus 2 SARS-CoV-2 uses the receptor angiotensin-converting enzyme (ACE) 2 for entry into target cells, and it was reported that both Angiotensin-converting enzyme inhibitors (ACEIs) and angiotensin receptor blockers (ARBs) could increase the mRNA expression of cardiac ACE2 receptors^20^. However, controversy about the novel use of Renin-angiotensin system (RAS) blockers has been raised amid this SARS-CoV-2 pandemic.

The explanation behind this controversy arises from the very fact that it shares the target receptor site with ACEIs and ARBs, which can cause the upregulation of ACE2 receptors^21^. ACE2 is additionally the notable cellular surface receptor and a necessary entry point for SARS-CoV-2 into the target cell^22^. As cardiovascular diseases and their therapy affects ACE2 levels, it plays an integral part in consideration of infectivity and outcomes of SARS-CoV-2. It needs to be imperatively determined whether treatment or disease-induced up-regulation of ACE2 impacts the trajectory of SARS-CoV-2^20^. ACEIs/ARBs are often used to treat hypertension, which is the most common comorbidity associated with SARS-CoV-2^23^. As there is no clinical evidence, major international cardiology societies recommend continuing the use of ACEIs and ARBs in SARS-CoV-2 patients^24^.

Due to limited literature on the influence of ACE inhibitors and ARBs in COVID-19 patients, we systematically reviewed the relevant medical literature. We performed a meta-analysis and meta-regression to investigate the association of ACEIs and ARBs used in COVID-19 and its effect on the mortality rate and severity of COVID-19.

### Methodology

We have presented this review according to the Preferred Systematic Reviews and Meta-Analysis Reporting Items guidelines for documenting analysis^25^.

### Search strategy

We searched WHO COVID-19 Global research database, Lit-COVID^26^, CDC Database of COVID-19 Research including PubMed, Embase, Scopus, Science Web, and Cochrane Central Controlled Trials Registry. The MedRxiv and SSNR preprint servers were also scanned. The searches were performed from December 2019 and revised till January 18^th^, 2021. The search approach and design can be found in Figure-1. Studies from all around the world were included, there were no language barriers. In an attempt to discover further eligible studies, we manually searched the reference lists of the included studies and the relevant literature. We also scanned the clinicaltrials.gov registry for completed, as well as in-progress randomized controlled trials (RCTs).

**Figure 1:**
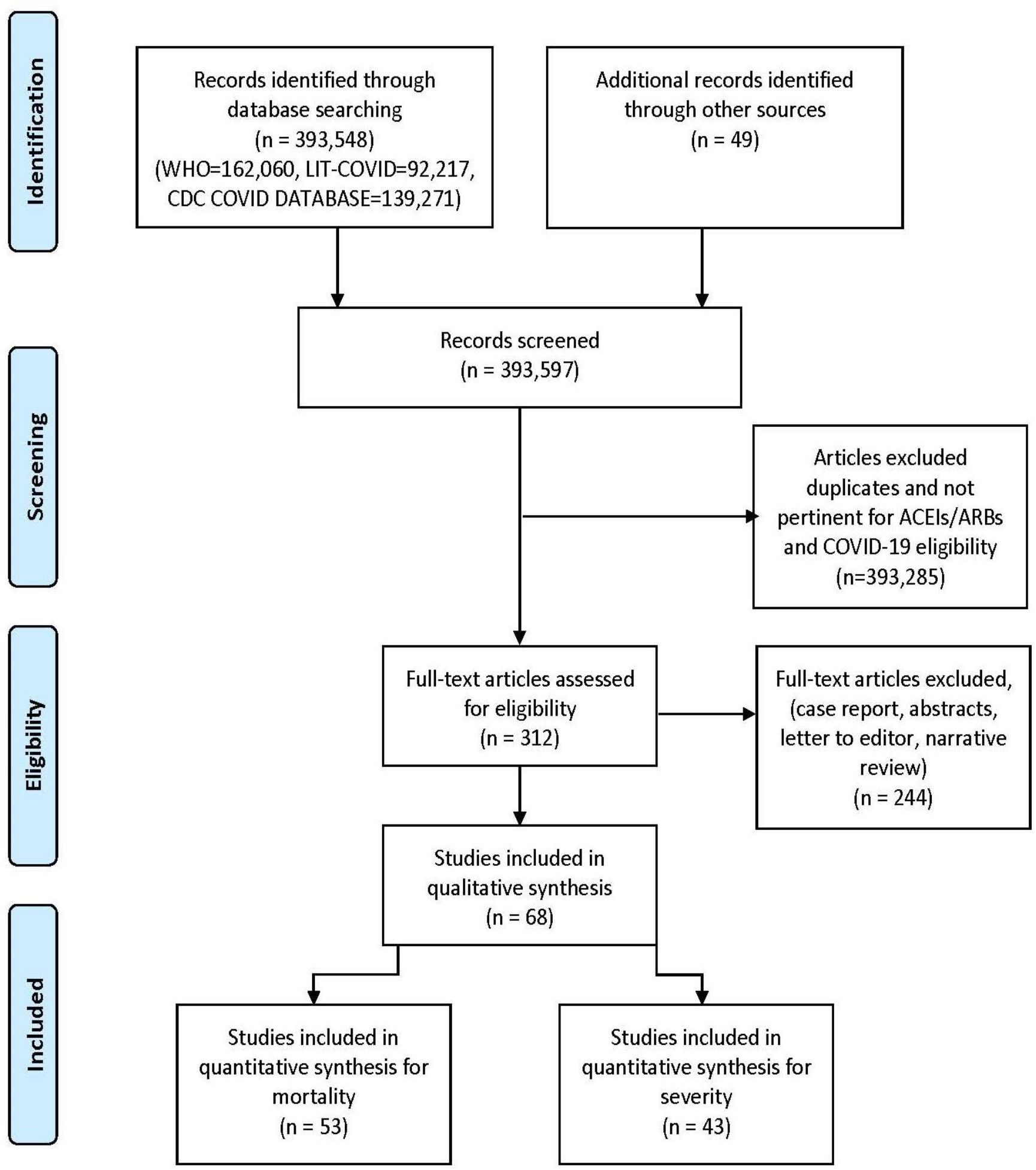
PRISMA flow diagram.

### Eligibility criteria

Criteria of eligibility: We included observational studies assessing the association between the use of the ACEI or ARB and at least one functional endpoint in COVID-19 positive patients diagnosed by RT-PCR. Case reports, narrative reviews, commentaries, and abstracts were excluded. The cases included Covid-19 positive patients on ACEI or ARB and the controls included the remaining COVID-19 positive patients.

### Study Selection

Three authors (RS, SSR, and HK) downloaded all articles from electronic search to EndNote X9^27^, as well as duplicates were eliminated. Titles and abstracts were autonomously evaluated by authors (AT, FA, HK, JM, KM, PG, RS, SA, and SSR) to identify and assess key articles. Further, authors (FA, HK, JM, KM, PG, RS, SA, and SSR) independently reviewed the entire manuscript and registered justification for the exclusion. Any discrepancies were addressed by arbitration.

### Outcome

All-cause mortality in the COVID-19 affected patient was the chief outcome, while severity of disease was the secondary outcome.

### Data Extraction

The authors (FA, HK, JM, KM, PG, RS, SA, and SSR), using a standardized data extraction method, extracted information from each survey independently; any conflict was resolved by consensus. The following dataset points were extracted: First author name, cases on ACEI-ARB, total COVID positive patients, country of study, study design, hypertension proportion, diabetes proportion, heart disease proportion, eligibility criteria, Median age, gender (female sex proportion), comorbidities, use of ACEIs or ARBs and primary and secondary outcomes (mortality and severity). Unadjusted and adjusted impact measurements were also extracted where appropriate.

### Statistical analysis

The meta-analysis specifically included longitudinal and cross-sectional studies comparing the effects of COVID-19 in subjects who were on ACEIs/ARBs at the time of infection with those who were not. Meta-analysis was performed first for studies reporting mortality of patients in both groups, followed by that for studies reporting severity of disease assuming independence of results for studies that reported both. Due to anticipated heterogeneity, summary statistics were calculated using a random-effects model. This model accounts for variability between studies as well as within studies. In all cases, meta-analyses were performed using the Mantel-Haenszel method for dichotomous data to estimate pooled odds ratios (OR) and statistical heterogeneity was assessed using Q value and I^2^ statistics. The meta-analysis and meta-regression was done with the Comprehensive Meta-Analysis software package (Biostat, Englewood, NJ, USA)^28^. We included the region of study in meta-regression model to find out whether Asian studies, which were dated earlier than studies from the rest of the world, contributed disproportionately to the significance of results. This helped rule out location and pipeline biases.

To explore differences between studies that might be expected to influence the effect size, we performed random effects (maximum likelihood method) univariate and multivariate meta-regression analyses. The potential sources of variability defined were median age of study sample, proportion of subjects of female sex, proportion of diabetics and proportion with heart diseases. Covariates were selected for further modeling if they significantly (*P* <0 .05) modified the association between mortality or severity in the COVID19 infected and treatment with ACEIs/ARBs. Two models were created, one for mortality and the other for severity of disease as outcomes. Subsequently, preselected covariates were included in a manual backward and stepwise multiple meta-regression analysis with *P* = 0.05 as a cutoff point for removal. *P* <0 .05 (*P* < 0.10 for heterogeneity) was considered statistically significant. All meta-analysis and meta-regression tests were 2-tailed.

## Results

The initial library search identified potentially relevant citations from WHO Global Research Database, CDC COVID-19 Research Articles Downloadable Database, and LitCovid PubMed database comprised of 393,597 articles. Subsequently, 393,285 articles were removed because of unclear evidence and non-relevance to the objective of the manuscript. Out of the remaining 312 articles, a total of 244 articles consisting of case reports, abstracts, letter to editor, and narrative reviews were excluded. Thus, 68 studies^29-96^ were included in their entirety as shown in Table 1. The PRISMA flow chart is shown in Figure 1.

**Table 1:** Study characteristics.

| First Author | Publication Status | Type of study | Outcome data | Country of Study | Median Age | Female Sex Proportion | Diabetes Proportion | Heart Diseases Proportion | Hypertension Proportion (in percentage) | High Severity (Case) | High Severity Sample size (case) | High Severity (Control) | High Severity Sample size (Control) | Mortality (Case) | Sample size (case) | Mortality (Control) | Sample size (control) |
| --- | --- | --- | --- | --- | --- | --- | --- | --- | --- | --- | --- | --- | --- | --- | --- | --- | --- |
| Alexandre et al. | peer-reviewed | Retrospective Cohort study | All-cause Mortality & Severity | UK | 61 | 39.76 | 28.24 | 17.57 | 48.7 | 17 | 117 | 12 | 230 | 17 | 117 | 12 | 230 |
| Anzola et al. | peer-reviewed | prospective study | Severity | Italy | 65 | 39 | 14 | 21 | 51 | 35 | 140 | 16 | 291 | N/A | N/A | N/A | N/A |
| Ashraf et al. | pre-print | Cross Sectional Cohort | Severity | Iran | 58 | 35.4 | 26 | 19 | 26 | 4 | 19 | 11 | 81 | N/A | N/A | N/A | N/A |
| Bae et al. | peer-reviewed | Cohort | All-cause Mortality & Severity | USA | 52 | 51.2 | 26.2 | 8.9 | 25.4 | 20 | 78 | 54 | 512 | 1 | 78 | 5 | 512 |
| Baker et al. | pre-print | Cohort | All-cause Mortality | UK | 75 | 46 | 26.6 | 20.6 | 42.1 | N/A | N/A | N/A | N/A | 17 | 78 | 63 | 233 |
| Banerjee et al. | peer-reviewed | Cohort | All-cause Mortality & Severity | UK | 61.5 | 42.8 | 43.8 | N/A | 100 | 1 | 1 | 1 | 6 | 1 | 1 | 0 | 6 |
| Bauer et al. | peer-reviewed | case-control study | All-cause Mortality | USA | 54.7 | 63 | 17 | 7 | 36 | N/A | N/A | N/A | N/A | 77 | 230 | 198 | 1219 |
| Bean et al. | peer-reviewed | Case-Control | All-cause Mortality & Severity | UK | 69.235 | 42.8 | 34.8 | 13.3 | 100 | 127 | 399 | 288 | 801 | 106 | 399 | 182 | 801 |
| Benelli et al. | pre-print | Cohort | All-cause Mortality | Italy | 66.8 | 33.4 | 16.3 | 22.6 | 46.95 | N/A | N/A | N/A | N/A | 25 | 110 | 47 | 301 |
| Braude et al. | peer-reviewed | multicenter observational study | All-cause Mortality | UK+ Italy | 74 | 40.9 | 27.1 | 21.8 | 51.5 | N/A | N/A | N/A | N/A | 106 | 392 | 257 | 979 |
| Bravi et al. | peer-reviewed | Case-Control | Severity | Italy | 58 | 52.3 | 12.1 | 16.1 | 33.9 | 267 | 450 | 69 | 93 | N/A | N/A | N/A | N/A |
| Cariou et al. | peer-reviewed | Cohort | All-cause Mortality & Severity | France | 69.8 | 35.1 | 88.5 | 11.6 | 77.2 | 232 | 737 | 150 | 580 | 92 | 737 | 48 | 580 |
| Cetinkal et al. | peer-reviewed | Retrospective single center study | All-cause Mortality & Severity | Turkey | 68.7 | 49.57 | 40.4 | 51 | 100 | 45 | 201 | 27 | 148 | 29 | 201 | 20 | 148 |
| Chaudri et al. | peer-reviewed | Single center Cohort study | All-cause Mortality & Severity | USA | 62 | 34 | 24.6 | 28.6 | 44.3 | 59 | 80 | 22 | 220 | 5 | 80 | 25 | 220 |
| Chen Ming et al. | pre-print | Case control | All-cause Mortality | China | 28.85 | 50.4 | 11.3 | 16.2 | 33.33 | N/A | N/A | N/A | N/A | 3 | 11 | 28 | 112 |
| Chen Yuchen et al. | peer-reviewed | Cohort | All-cause Mortality | China | 67.25 | 33 | 100 | N/A | 100 | N/A | N/A | N/A | N/A | 4 | 32 | 10 | 39 |
| Choi et al. | pre-print | Case Control | All-cause Mortality & Severity | South Korea | 66.5 | 57.2 | 44.9 | N/A | 100 | 34 | 892 | 55 | 625 | 42 | 892 | 69 | 625 |
| Conversano et al. | peer-reviewed | Cohort | All-cause Mortality | Italy | 65 | 31.4 | 14.6 | 4.7 | 50.26 | N/A | N/A | N/A | N/A | 48 | 69 | 101 | 122 |
| De Spiegele er et al. | peer-reviewed | Cohort | Severity | Europe | 85.9 | 67 | 18.1 | N/A | 25.3 | 6 | 30 | 31 | 124 | N/A | N/A | N/A | N/A |
| Covino et al. | peer-reviewed | Retrospective study | All-cause Mortality & Severity | Austral ia | 74 | 34.3 | 13.2 | 42.1 | 100 | 38 | 51 | 82 | 115 | 58 | 111 | 22 | 55 |
| Desai et al. | peer-reviewed | retrospective study | All-cause Mortality | Italy | 64.8 | 33.9 | 20 | 27.1 | 43.1 | N/A | N/A | N/A | N/A | 49 | 154 | 72 | 421 |
| Felice et al. | peer-reviewed | Cohort | All-cause Mortality & Severity | Italy | 72 | 35.4 | 25.5 | 18 | 100 | 21 | 82 | 25 | 51 | 15 | 82 | 18 | 51 |
| Zhou feng et al. | peer-reviewed | Cohort | All-cause Mortality | China | 66 | 49.9 | N/A | N/A | 100 | N/A | N/A | N/A | N/A | 70 | 906 | 272 | 1812 |
| Fosbol et al. | peer-reviewed | Cohort | All-cause Mortality & Severity | Denma rk | 62 | 52 | 9 | 8.5 | 18.8 | 203 | 895 | 373 | 3585 | 181 | 895 | 297 | 3585 |
| Golpe et al. | peer-reviewed | Cohort | Severity | Spain | 70.4 | 54.1 | 9.8 | N/A | 29.1 | 48 | 69 | 73 | 88 | N/A | N/A | N/A | N/A |
| Guner et al. | peer-reviewed | Cohort | Severity | Turkey | 50.6 | 40.5 | 13.5 | 23.6 | 23.4 | 21 | 65 | 29 | 167 | N/A | N/A | N/A | N/A |
| Genet et al. | peer-reviewed | Retrospective Observational study | All-cause Mortality & Severity | USA | 86.3 | 36.19 | 10.45 | 12.6 | 33.5 | 14 | 63 | 52 | 138 | 14 | 63 | 52 | 138 |
| Guo et al. | peer-reviewed | Cross Sectional | All-cause Mortality | China | 58 | 51.3 | 15 | 15.5 | 32.62 | N/A | N/A | N/A | N/A | 7 | 19 | 36 | 168 |
| Hakeam et al. | peer-reviewed | Multicenter prospective Cohort | All-cause Mortality & Severity | Saudi Arabia | 60.8 | 40.5 | 63.3 | 32.2 | 93.7 | 69 | 245 | 33 | 93 | 15 | 69 | 7 | 33 |
| Huh et al. | peer-reviewed | retrospective cohort study | Severity | Korea | 47.1 | 59.54 | 16.06 | 7.83 | 21.47 | 248 | 877 | 630 | 6464 | N/A | N/A | N/A | N/A |
| Huang et al. | peer-reviewed | Cohort | All-cause Mortality | China | 60.185 | 55 | 8 | 2 | 100 | N/A | N/A | N/A | N/A | 0 | 20 | 3 | 30 |
| Kim Ju Hwan et al. | peer-reviewed | Retrospective Cohort study | All-cause Mortality & Severity | South Korea | 62.8 | 49.79 | 62.69 | 53.89 | 22.6 | 23 | 628 | 28 | 608 | 23 | 628 | 28 | 608 |
| Ip et al. | pre-print | Cross Sectional | All-cause Mortality | USA | N/A | N/A | N/A | N/A | 52.5 | N/A | N/A | N/A | N/A | 137 | 460 | 262 | 669 |
| Jurado et al. | pre-print | Cross Sectional | Severity | Spain | 63.2 | 40.6 | 23.8 | N/A | 52.4 | 56 | 92 | 135 | 198 | N/A | N/A | N/A | N/A |
| Jung et al. | peer-reviewed | Cohort | All-cause Mortality | South Korea | 44.6 | 56 | 17 | 5 | 22.34 | N/A | N/A | N/A | N/A | 33 | 377 | 51 | 1577 |
| Kim Lindsay et al. | peer-reviewed | Cohort | All-cause Mortality | USA | 62 | 46.8 | 32.9 | 34.6 | 57.32 | N/A | N/A | N/A | N/A | 105 | 573 | 530 | 3214 |
| Lafaurie et al. | peer-reviewed | Cohort study | All-cause Mortality & Severity | UK | 74 | 77.56 | 12.92 | 12.9 | 38.78 | 36 | 73 | 14 | 36 | 9 | 73 | 6 | 36 |
| Lee et al. | pre-print | Cohort | All-cause Mortality | South Korea | 44.36 | 77.56 | 17 | 6.7 | 19 | N/A | N/A | N/A | N/A | 50 | 977 | 62 | 7289 |
| Li et al. | peer-reviewed | Cross Sectional | All-cause Mortality & Severity | China | 66 | 47.8 | 35 | 35.07 | 30.73 | 57 | 115 | 116 | 247 | 21 | 115 | 56 | 247 |
| Liu et al. | pre-print | Cross Sectional | Severity | China | 65.2 | 44.9 | N/A | N/A | 15.26 | 7 | 22 | 31 | 56 | N/A | N/A | N/A | N/A |
| **Mancia et al.** | peer-reviewed | Cross Sectional | Severity | Italy | 68 | 36.7 | N/A | 30.1 | 58 | 364 | 2896 | 253 | 3376 | N/A | N/A | N/A | N/A |
| **Lim et al.** | peer-reviewed | Retrospective Cohort study | All-cause Mortality & Severity | South Korea | 67 | 46.15 | 25.38 | 10 | 40 | 14 | 30 | 22 | 100 | 14 | 30 | 22 | 100 |
| **Mehta et al.** | peer-reviewed | Cohort | All-cause Mortality | USA | 58.5 | 49.9 | 41.1 | 19.9 | 11.6 | N/A | N/A | N/A | N/A | 8 | 211 | 34 | 1494 |
| **Meng et al.** | peer-reviewed | Cross Sectional | All-cause Mortality & Severity | China | 64.5 | 42.8 | 30.95 | N/A | 10 | 4 | 17 | 12 | 25 | 0 | 17 | 1 | 25 |
| **Lopez-Otero et al.** | peer-reviewed | Cohort | All-cause Mortality | Spain | 64.05 | 56 | 13 | 21.2 | 100 | N/A | N/A | N/A | N/A | 11 | 211 | 27 | 755 |
| **Oussalah et al.** | peer-reviewed | retrospective longitudinal cohort analysis | All-cause Mortality & Severity | France | 65 | 39 | 29 | 29 | 50 | 20 | 44 | 34 | 105 | 10 | 43 | 9 | 104 |
| **Peng et al.** | peer-reviewed | Cross Sectional | All-cause Mortality & Severity | China | 31 | 52.68 | 26 | 66.3 | 82.14 | 3 | 22 | 13 | 90 | 4 | 22 | 13 | 90 |
| **Rezel-Potts et al.** | peer-reviewed | case control study with additional cohort analysis | All-cause Mortality | UK | 62 | 60 | 13 | 14 | 26.8 | N/A | N/A | N/A | N/A | 254 | 2712 | 667 | 14154 |
| **Reynolds et al.** | peer-reviewed | Cross Sectional | Severity | USA | 32 | 58.5 | 39.7 | 16.1 | 34.59 | 587 | 2401 | 608 | 2393 | N/A | N/A | N/A | N/A |
| **Rhee et al.** | pre-print | Cohort | Severity | South Korea | 62.375 | 46.5 | N/A | 18.5 | 68.02 | 13 | 327 | 21 | 505 | N/A | N/A | N/A | N/A |
| **Rentsch et al.** | pre-print | Cohort | All-cause Mortality | USA | 66.1 | 4.6 | 44.4 | 27.9 | 71.96 | N/A | N/A | N/A | N/A | 11 | 255 | 6 | 324 |
| **Richardson et al.** | peer-reviewed | Cross Sectional | All-cause Mortality & Severity | USA | 63 | 39.7 | 59.8 | 33.8 | 53.08 | 87 | 413 | 141 | 953 | 130 | 413 | 254 | 953 |
| **Rodilla et al.** | peer-reviewed | Cross-sectional, observational, retrospective multicenter study, | All-cause Mortality | Spain | 67.5 | 42.6 | 19.1 | 4 | 50.9 | N/A | N/A | N/A | N/A | 1180 | 4238 | 1452 | 7988 |
| **Rosenthal et al.** | peer-reviewed | retrospective cohort study | All-cause Mortality | USA | 57 | 50.1 | 27.9 | 22.09 | 46.7 | N/A | N/A | N/A | N/A | 345 | 3162 | 7010 | 34545 |
| **Rossi et al.** | pre-print | Cohort | All-cause Mortality | Italy | 31.6 | 49.9 | 12 | 5.8 | 16.2 | N/A | N/A | N/A | N/A | 108 | 818 | 109 | 1835 |
| **Soleimani et al.** | peer-reviewed | Retrospective Observational study | All-cause Mortality & Severity | USA | 66.4 | 16.5 | 0.18 | 13.99 | 39.93 | 91 | 122 | 91 | 132 | 33 | 122 | 35 | 132 |
| **Tan et al.** | peer-reviewed | Case Control | All-cause Mortality & Severity | China | 67.25 | 49 | 28 | 18 | 100 | 27 | 31 | 60 | 69 | 0 | 31 | 11 | 69 |
| **W Lam et al.** | peer-reviewed | Retrospective single center study | All-cause Mortality & Severity | USA | 70.5 | 9.89 | 8.96 | 8.24 | 31.37 | 65 | 335 | 55 | 279 | 58 | 335 | 62 | 279 |
| **Wang et al.** | peer-reviewed | Cross Sectional | All-cause Mortality | China | 64 | 48 | 18.6 | 11.6 | 41.86 | N/A | N/A | N/A | N/A | 30 | 62 | 103 | 282 |
| **Zhou Xian et al.** | peer-reviewed | Case-Control | All-cause Mortality | China | 57.7 | 45.5 | 10 | 9.1 | 32.7 | N/A | N/A | N/A | N/A | 2 | 15 | 5 | 21 |
| **Yan et al.** | pre-print | Cohort | Severity | China | 48.75 | 48.9 | 10 | 2.62 | 22.46 | 52 | 256 | 76 | 354 | N/A | N/A | N/A | N/A |
| **Liu Xiulan et al.** | peer-reviewed | retrospective single-center case series | All-cause Mortality & Severity | China | 65.39 | 53.5 | 22 | 16 | 100 | 31 | 74 | 38 | 83 | 5 | 74 | 1 | 83 |
| Yang et al. | pre-print | Cohort | All-cause Mortality & Severity | China | 66.5 | 50.8 | 21.9 | 14 | 50.5 | 15 | 42 | 35 | 83 | 2 | 43 | 11 | 83 |
| Yun Feng et al. | peer-reviewed | Case-Control | Severity | China | 53 | 43.1 | 10.3 | 8 | 25.8 | 4 | 33 | 36 | 80 | N/A | N/A | N/A | N/A |
| Zeng et al. | pre-print | Cross Sectional | All-cause Mortality & Severity | China | 66.5 | 50.6 | 56 | 41.3 | 27.37 | 15 | 28 | 15 | 47 | 2 | 28 | 5 | 47 |
| Zhang et al. | peer-reviewed | Cohort | All-cause Mortality | China | 64 | 46 | 21.3 | 29.09 | 100 | N/A | N/A | N/A | N/A | 7 | 188 | 92 | 940 |
| Zhichao Feng et al. | pre-print | Cohort | Severity | China | 47 | 49.6 | 8 | 4 | 14.5 | 1 | 16 | 16 | 49 | N/A | N/A | N/A | N/A |
| Zhong et al. | peer-reviewed | Retrospective Observational study | All-cause Mortality & Severity | UK | 66.3 | 55.5 | 32.5 | 16.6 | 100 | 6 | 37 | 15 | 89 | 6 | 37 | 15 | 89 |

### Study characteristics of included studies

A total of fifty three studies^29-81^ were included for meta-analysis for the primary outcome i.e. mortality. In total, these consisted of 112,468 subjects with 16,363 mortality events. Median age for all studies was 64.9 (60.9-67.2) with average 47.3% females (Table 1). Of the comorbidities considered, 25.4% were diabetics, 16.6% had heart diseases overall. Similarly, a total of forty three studies^29,31,33,36-38,41,43,45-47,49,53,55-57,59-61,64-66,69,73,74,76,77,79,82-96^ were included for meta-analysis for the secondary outcome i.e. severity of disease. These had a combined sample size of 37,914 with 6,985 patients reaching the endpoint of high disease severity. The median age was 65.2 (60.8-68.7) and 46.2% were females, 25.6% were diabetics, 17.1% had heart diseases overall in this cohort (Table 1).

### Meta-analysis for mortality outcome

Meta-analysis findings showed that being on ACEIs/ARBs did not have an association with mortality from COVID 19 infections compared to not being on ACEIs/ARBs (OR=1.17, 95% CI 0.94 to 1.45, p= 0.15). Heterogeneity was very high with I^2^ = 93.2% (Figure 2).

**Figure 2:**
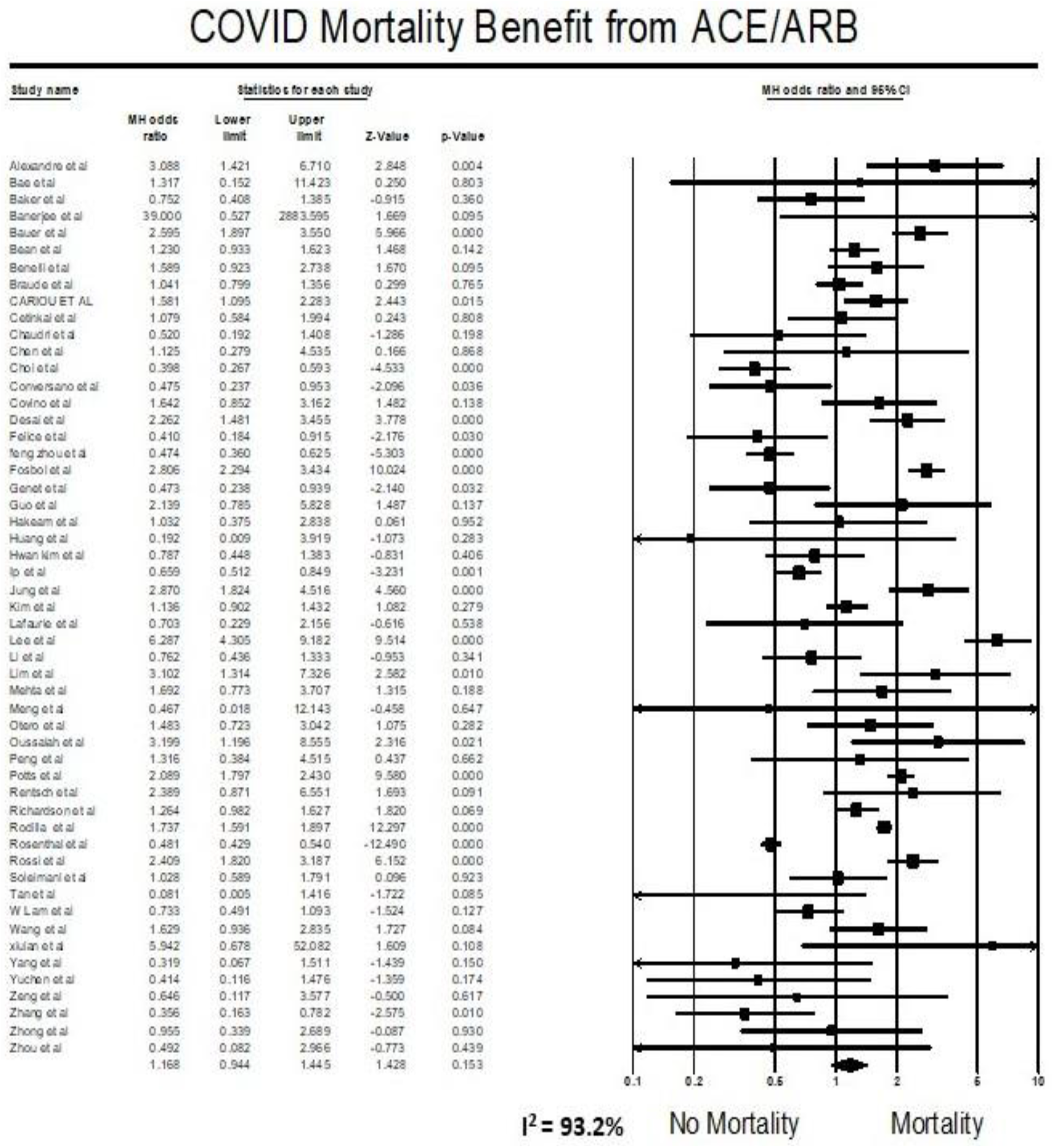
Forest plot for association of ACE/ARBs on mortality in COVID19 patients.

### Meta-analysis for severity outcome

Findings from the meta-analysis showed that being on ACEI/ARBs did not have an association with severity from COVID 19 infections compared to not being on ACEIs/ARBs (OR=1.18, 95% CI 0.94 to 1.48, p= 0.15). Heterogeneity was very high with I^2^ = 91.1% (Figure 3).

**Figure 3:**
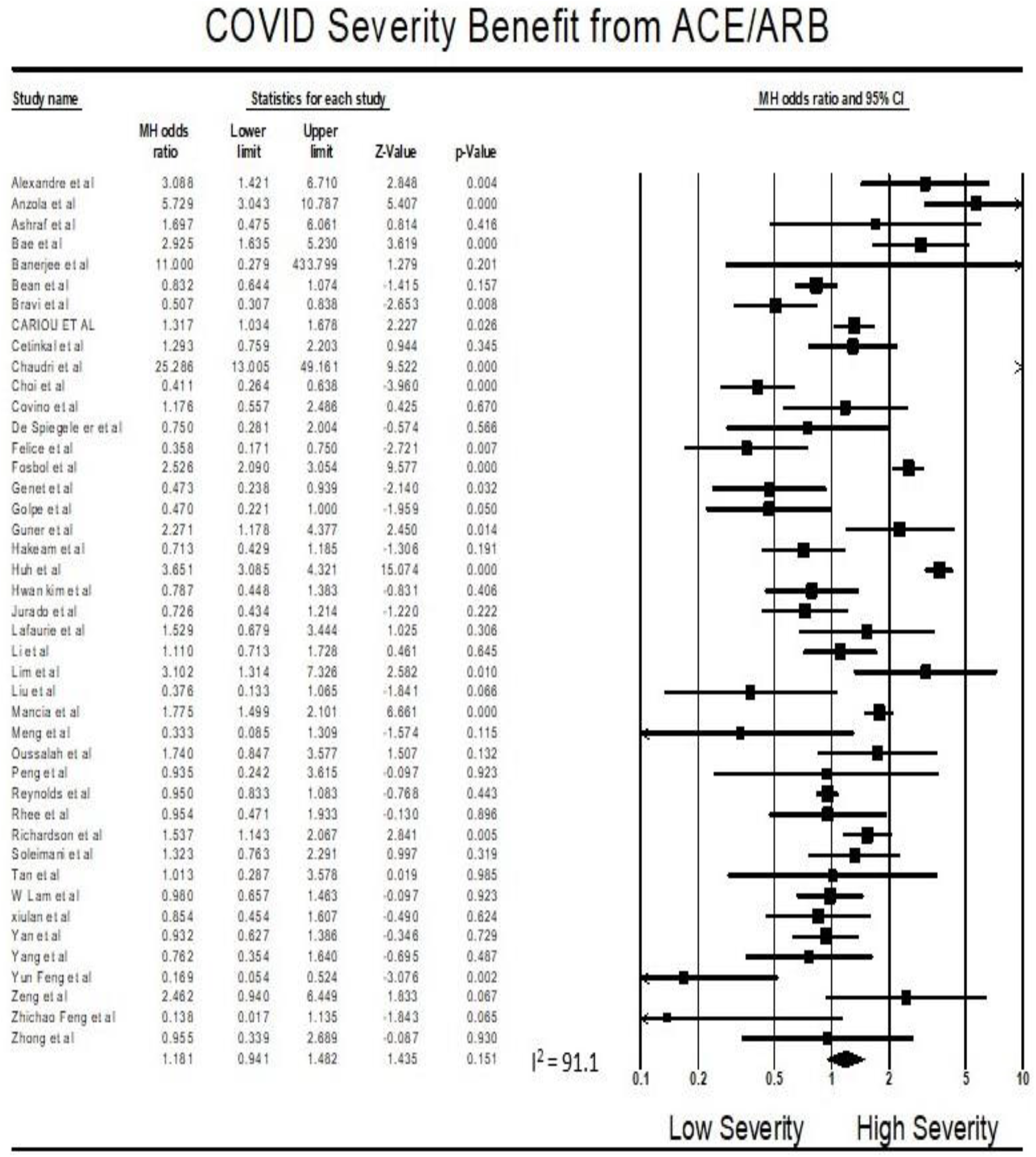
Forest plot for association of ACE/ARBs on severity in COVID19 patients.

### Multivariate meta-regression model for mortality outcome

Multivariate meta-regression performed to explain variations in association between mortality and being on ACEIs/ARBs revealed; age, female gender, proportion of heart diseases in included studies covariates to be significant together and explained R^2^=36% of the between study heterogeneity in mortality. Figure 4 shows the resulting equation and individual covariate effect graphs.

**Figure 4:**
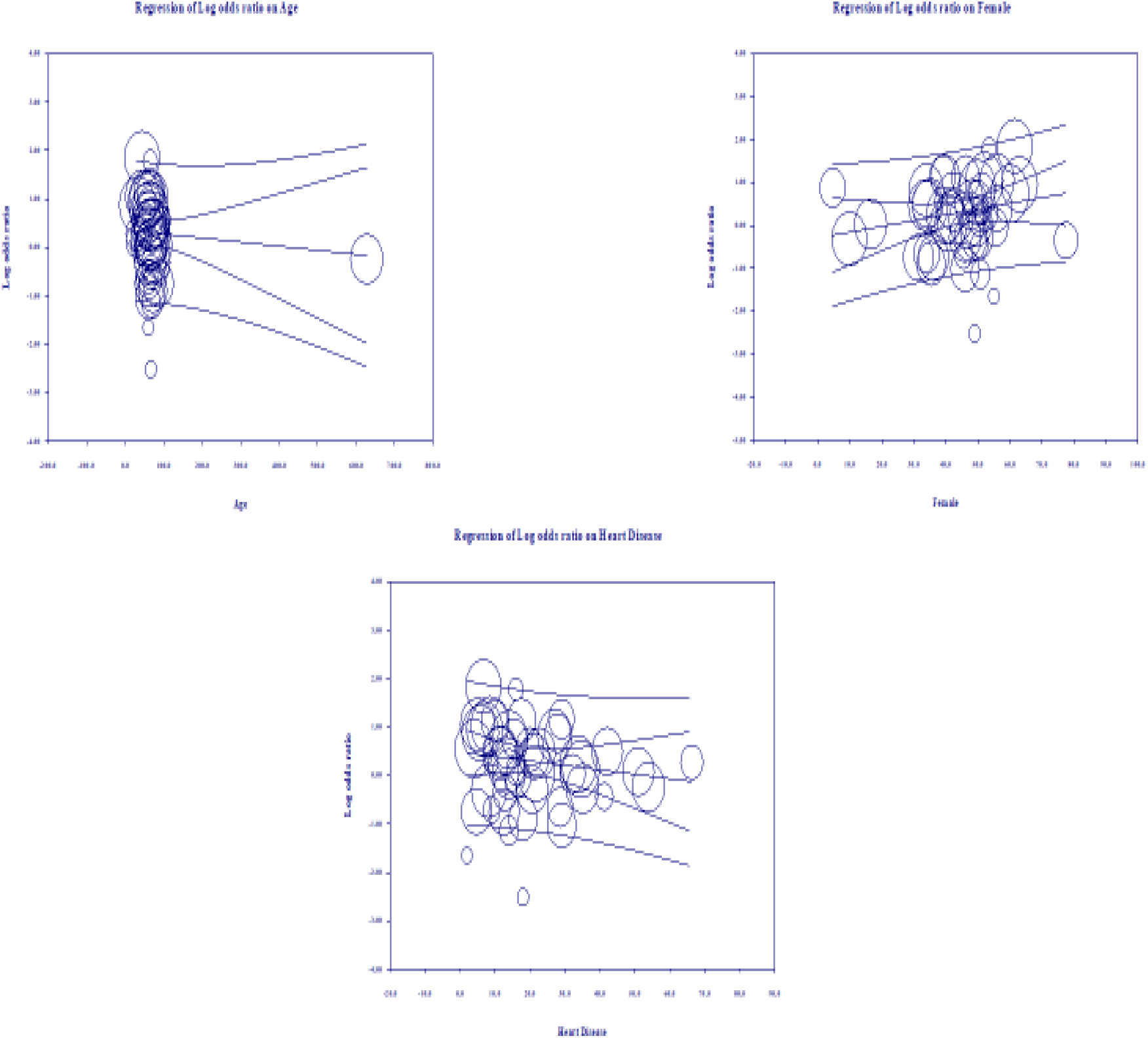
Meta-regression results for covariates significantly influencing mortality (R^2^=36)

### Multivariate meta-regression model for severity outcome

Multivariate meta-regression performed to explain variations in association between severity and being on ACEIs/ARBs revealed age, proportion with diabetes, heart disease and country of studies covariates to be significant together. These covariates together explained R^2^= 8% of the study heterogeneity in severity. Figure 5 shows the resulting equation and individual covariate effect graphs.

**Figure 5:**
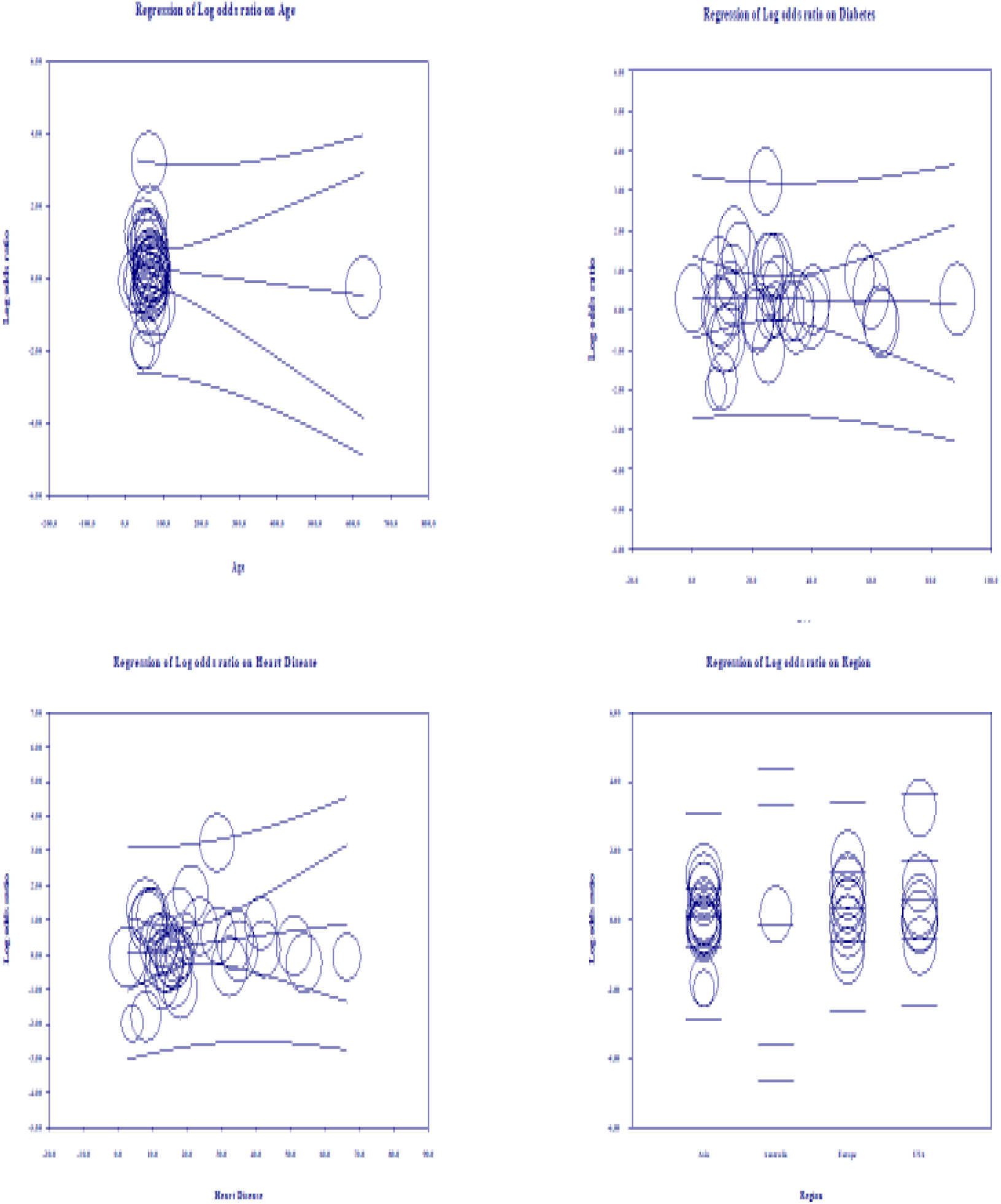
Meta-regression results for covariates significantly influencing severity (R^2^=8%)

### Risk of bias assessment

The Newcastle-Ottawa (NOS) scale^97^ was used for measuring the risk of bias in cohort and case-control studies (Table 2A & 2B). The following classes were rated per study: low bias risk (9 points), moderate bias risk (5-7 points), and high bias risk (0-4 items). For a cross-sectional study, we used the modified version of NOS, assigning the study in the following groups: Low risk of bias (8-10), moderate risk (5-7), high risk of bias (0-4). Three reviewers (RS, SSR, and PG) evaluated the likelihood of bias independently, and any conflict was resolved by consensus.

**Table 2:**
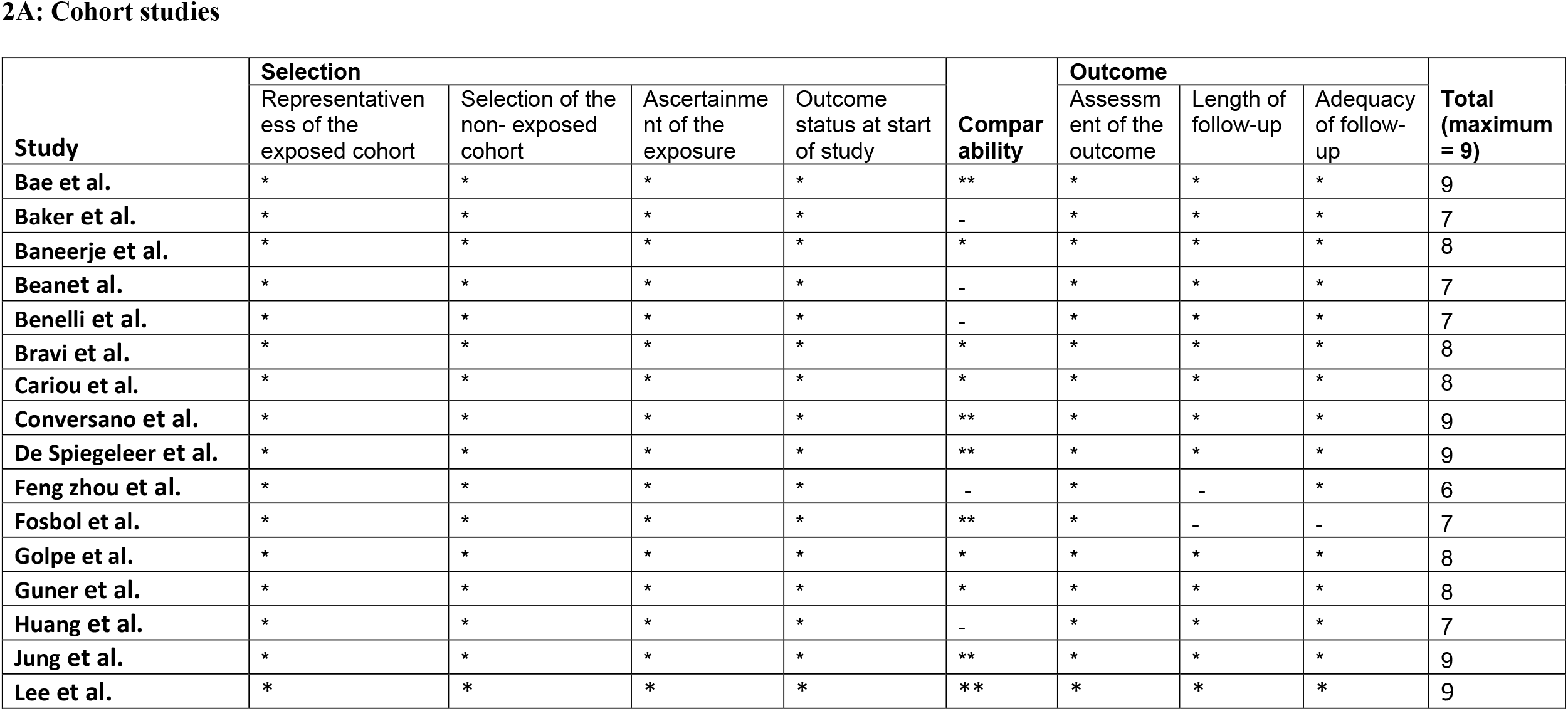
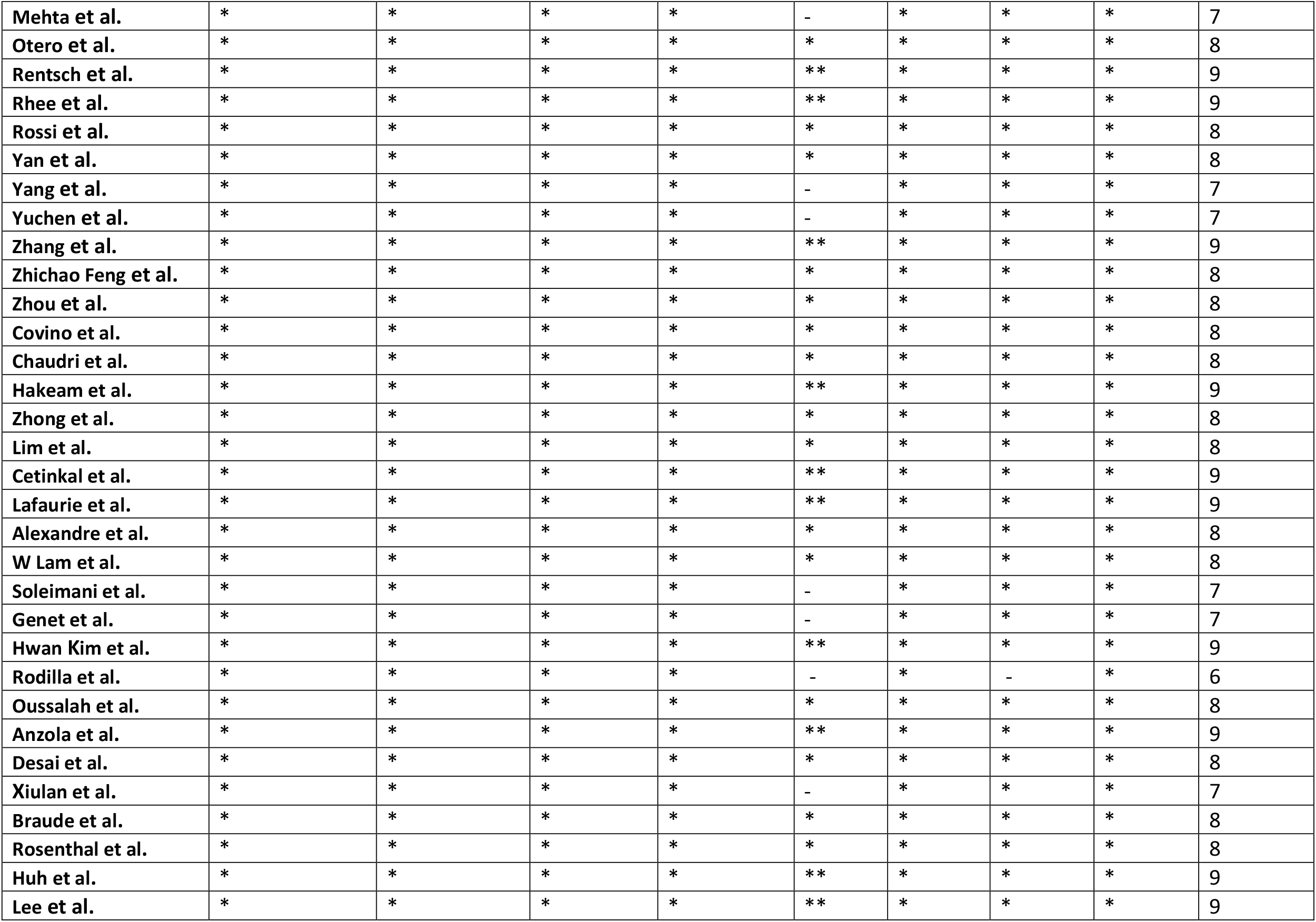
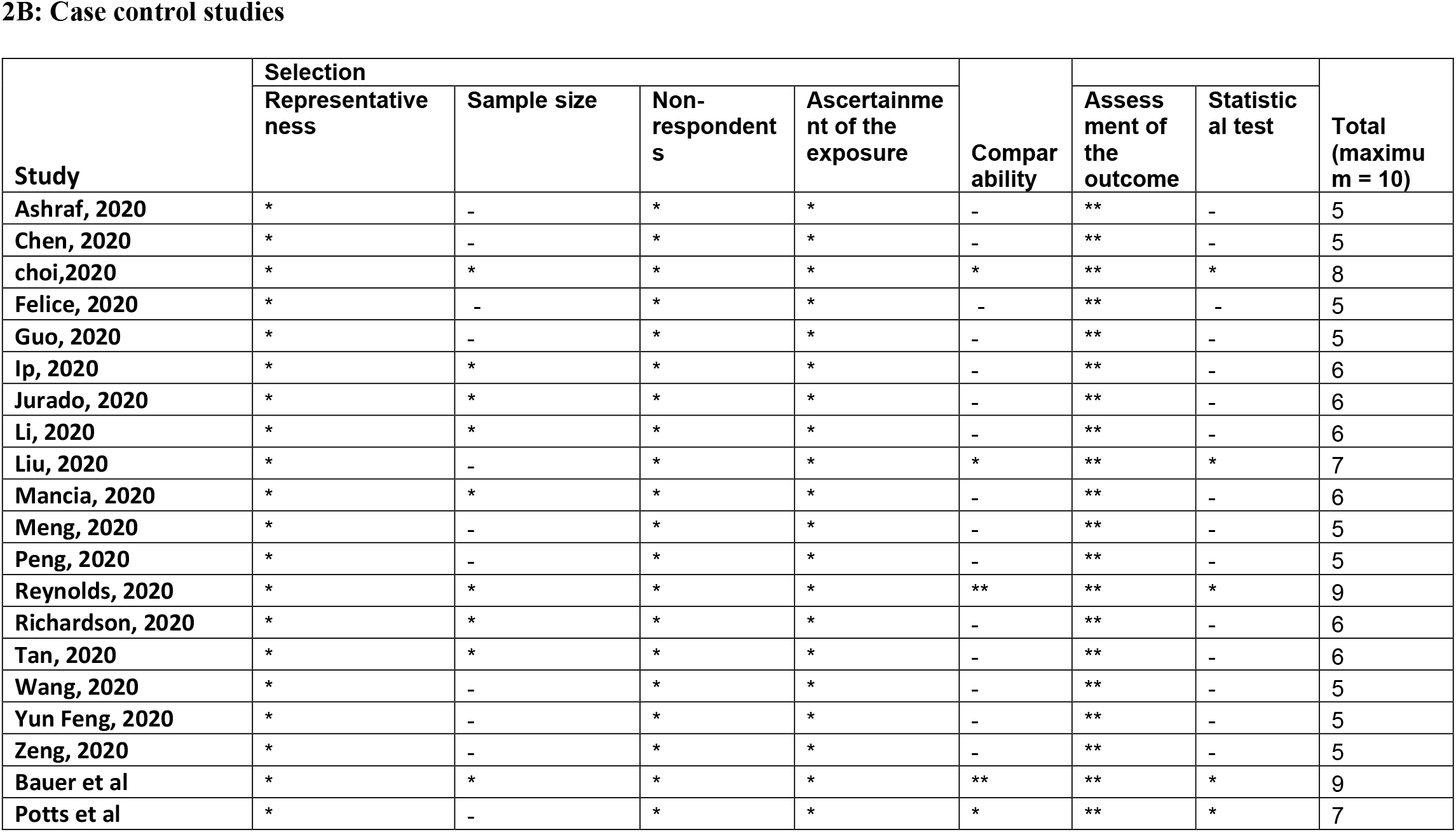
New Castle Ottawa Scale.

2A: Cohort studies
| Study | Selection |  |  |  | Comparability | Outcome |  |  | Total (maximum = 9) |
| --- | --- | --- | --- | --- | --- | --- | --- | --- | --- |
|  | Representativeness of the exposed cohort | Selection of the non- exposed cohort | Ascertainment of the exposure | Outcome status at start of study |  | Assessment of the outcome | Length of follow-up | Adequacy of follow-up |  |
| Bae et al. | * | * | * | * | ** | * | * | * | 9 |
| Baker et al. | * | * | * | * | - | * | * | * | 7 |
| Baneerje et al. | * | * | * | * | * | * | * | * | 8 |
| Beanet al. | * | * | * | * | - | * | * | * | 7 |
| Benelli et al. | * | * | * | * | - | * | * | * | 7 |
| Bravi et al. | * | * | * | * | * | * | * | * | 8 |
| Cariou et al. | * | * | * | * | * | * | * | * | 8 |
| Conversano et al. | * | * | * | * | ** | * | * | * | 9 |
| De Spiegeleer et al. | * | * | * | * | ** | * | * | * | 9 |
| Feng zhou et al. | * | * | * | * | - | * | - | * | 6 |
| Fosbol et al. | * | * | * | * | ** | * | - | - | 7 |
| Golpe et al. | * | * | * | * | * | * | * | * | 8 |
| Guner et al. | * | * | * | * | * | * | * | * | 8 |
| Huang et al. | * | * | * | * | - | * | * | * | 7 |
| Jung et al. | * | * | * | * | ** | * | * | * | 9 |
| Lee et al. | * | * | * | * | ** | * | * | * | 9 |
| Mehta et al. | * | * | * | * | - | * | * | * | 7 |
| Otero et al. | * | * | * | * | * | * | * | * | 8 |
| Rentsch et al. | * | * | * | * | ** | * | * | * | 9 |
| Rhee et al. | * | * | * | * | ** | * | * | * | 9 |
| Rossi et al. | * | * | * | * | * | * | * | * | 8 |
| Yan et al. | * | * | * | * | * | * | * | * | 8 |
| Yang et al. | * | * | * | * | - | * | * | * | 7 |
| Yuchen et al. | * | * | * | * | - | * | * | * | 7 |
| Zhang et al. | * | * | * | * | ** | * | * | * | 9 |
| Zhichao Feng et al. | * | * | * | * | * | * | * | * | 8 |
| Zhou et al. | * | * | * | * | * | * | * | * | 8 |
| Covino et al. | * | * | * | * | * | * | * | * | 8 |
| Chaudri et al. | * | * | * | * | * | * | * | * | 8 |
| Hakeam et al. | * | * | * | * | ** | * | * | * | 9 |
| Zhong et al. | * | * | * | * | * | * | * | * | 8 |
| Lim et al. | * | * | * | * | * | * | * | * | 8 |
| Cetinkal et al. | * | * | * | * | ** | * | * | * | 9 |
| Lafaurie et al. | * | * | * | * | ** | * | * | * | 9 |
| Alexandre et al. | * | * | * | * | * | * | * | * | 8 |
| W Lam et al. | * | * | * | * | * | * | * | * | 8 |
| Soleimani et al. | * | * | * | * | - | * | * | * | 7 |
| Genet et al. | * | * | * | * | - | * | * | * | 7 |
| Hwan Kim et al. | * | * | * | * | ** | * | * | * | 9 |
| Rodilla et al. | * | * | * | * | - | * | - | * | 6 |
| Oussalah et al. | * | * | * | * | * | * | * | * | 8 |
| Anzola et al. | * | * | * | * | ** | * | * | * | 9 |
| Desai et al. | * | * | * | * | * | * | * | * | 8 |
| Xiulan et al. | * | * | * | * | - | * | * | * | 7 |
| Braude et al. | * | * | * | * | * | * | * | * | 8 |
| Rosenthal et al. | * | * | * | * | * | * | * | * | 8 |
| Huh et al. | * | * | * | * | ** | * | * | * | 9 |
| Lee et al. | * | * | * | * | ** | * | * | * | 9 |

**2B: Case control studies**
| Study | Selection |  |  |  | Comparability |  |  | Total (maximum = 10) |
| --- | --- | --- | --- | --- | --- | --- | --- | --- |
|  | Representativeness | Sample size | Non-respondents | Ascertainment of the exposure |  | Assessment of the outcome | Statistical test |  |
| Ashraf, 2020 | * | - | * | * | - | ** | - | 5 |
| Chen, 2020 | * | - | * | * | - | ** | - | 5 |
| choi,2020 | * | * | * | * | * | ** | * | 8 |
| Felice, 2020 | * | - | * | * | - | ** | - | 5 |
| Guo, 2020 | * | - | * | * | - | ** | - | 5 |
| Ip, 2020 | * | * | * | * | - | ** | - | 6 |
| Jurado, 2020 | * | * | * | * | - | ** | - | 6 |
| Li, 2020 | * | * | * | * | - | ** | - | 6 |
| Liu, 2020 | * | - | * | * | * | ** | * | 7 |
| Mancia, 2020 | * | * | * | * | - | ** | - | 6 |
| Meng, 2020 | * | - | * | * | - | ** | - | 5 |
| Peng, 2020 | * | - | * | * | - | ** | - | 5 |
| Reynolds, 2020 | * | * | * | * | ** | ** | * | 9 |
| Richardson, 2020 | * | * | * | * | - | ** | - | 6 |
| Tan, 2020 | * | * | * | * | - | ** | - | 6 |
| Wang, 2020 | * | - | * | * | - | ** | - | 5 |
| Yun Feng, 2020 | * | - | * | * | - | ** | - | 5 |
| Zeng, 2020 | * | - | * | * | - | ** | - | 5 |
| Bauer et al | * | * | * | * | ** | ** | * | 9 |
| Potts et al | * | - | * | * | * | ** | * | 7 |

### Publication Bias

Visual inspection of the standard error plots for the mortality analysis (Figure 6A) suggests symmetry without an underrepresentation of studies of any precision but indicated underrepresentation of studies with smaller effect sizes. Classic fail-safe N analysis computed taking alpha at 0.05 put the number of missing studies at 5. Corroborating inspection findings, in Egger’s regression test the null hypothesis of no small study effects was not rejected at P<0.05 (estimated bias coefficient = −0.28 ± 0.76 SE).

**Figure 6:**
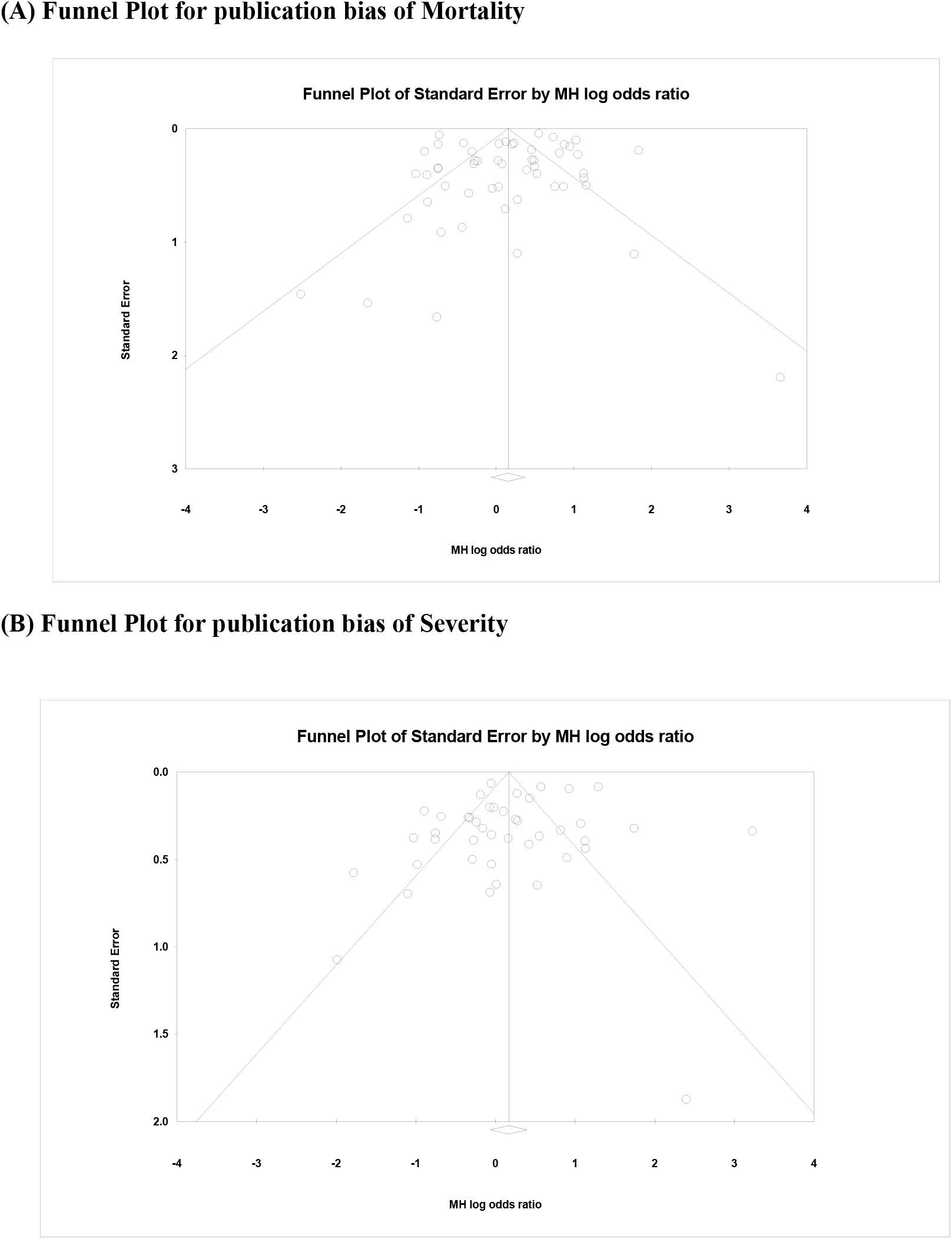
Funnel plots for publication bias of Mortality (A) and Severity (B) models:

Similarly, visual inspection of the standard error plots for the severity analysis also (Figure 6B) suggest symmetry without an underrepresentation of studies of any precision but indicated underrepresentation of studies with smaller effect sizes. Classic fail-safe N analysis computed taking alpha at 0.05 put the number of missing studies at 8. However, in Egger’s regression test the null hypothesis of no small study effects was rejected at P<0.05 (estimated bias coefficient = −1.14 ± 0.81SE).

## Discussion

Based on our meta-analysis consisting of cross-sectional, case-control, and cohort studies, the use of ACEIs and ARBs was neither associated with increased all-cause mortality nor with increased severity of disease progression in COVID-19 patients. Multivariate meta-regression for the mortality model demonstrated that 36% of study variations could be explained by differences in age, female gender, proportion of heart diseases in the study samples. Multivariate meta-regression for the severity model demonstrated that 8% of study variations could be explained by differences in age, proportion of diabetes, heart diseases and country of studies in the study samples. This finding is valuable as association between ACEIs and ARBs use and outcome in COVID-19 patients has been inconclusive so far. To our knowledge, this is the first meta-regression, and the largest meta-analysis to evaluate the role of ACEIs/ARBs as an antihypertensive regimen in hospitalized patients with COVID-19.

The effect of ACEIs/ARBs use on COVID-19 patients has been a controversial topic since the beginning of this pandemic, and some studies even have interposed a risk of taking ACEIs/ARBs using data from previous coronavirus outbreaks and preclinical studies^98^. Previously published systematic reviews, suggested a lower mortality (25-43%) in patients with hypertension hospitalized for COVID19^99-101^. Furthermore, a large-scale retrospective study demonstrated that in-hospital use of ACEIs/ARBs was associated with a lower risk of 28-day death among hospitalized patients with COVID-19 and coexisting hypertension (adjusted HR 0.32, 95% CI 0.15 to 0.66)^78^. These data suggested that patients with hypertension might obtain benefits from taking ACEIs/ARBs compared with the non-ACEIs/ARBs in the setting of COVID-19 and support the hypothesis that a drug that diminishes angiotensin-2 activity, such as ACEIs and/or ARBs, can reduce the deadliness of inflammation associated injury in COVID-19. Our result trends in contraindicating the above study results but did not reach statistical significance. Our meta-analysis suggests that the use of ACEIs/ARBs neither increase nor decrease mortality in COVID-19 patients (figure 2). In addition to what is reported in published studies, our systematic review added the most recent studies, and had the largest sample size (53 studies, 112,468 patients).

The main strength of our analysis is the large sample size along with a robust and comprehensive search. The large sample size enables the precision and reliability of risk estimates. Additionally, further meta-regression was performed to adjust for confounding factors. However, despite all the strengths, there are still certain limitations. The major limitation of the meta-regression is the presence of unknown confounders. Multiple previous studies have reported that gender, age, smoking history, and presence of diabetes influence COVID-19 results. Even though these confounders are reported in most of the included studies, further studies focusing on the adjustment of confounders are necessary. We included studies from the medRxiv.org databases and other preprint database which did not go through peer review at that time. We considered this as a limitation, as peer reviewers could catch more deficiencies in reporting methods and other details. However, it was anticipated that majority of these studies would be peer-reviewed. Third, the use of ACE/ARB has been via medical record review which could be less reliable. Finally, the definition of COVID-19 severity and outcomes were not uniform among the included studies.

In conclusion, meta-regression analysis suggest that the use of ACEIs/ARBs in patients with COVID-19 is not associated with increased mortality or increased severity. However, multivariate meta-regression model for mortality indicated that 36% study variation could be due to differences in age, female gender, proportion of heart diseases in the study samples. Similarly, multivariate meta-regression for the severity model exhibited that 8% of study variations could be explained by differences in age, proportion of diabetes, heart disease and country of studies in the study samples. Larger observational studies^102-105^ and clinical trials are warranted to confirm these findings. Providers should continue to manage patient hypertension as per current treatment guidelines^106,107^ and clinical judgement.

## Data Availability

We performed a meta-analysis from already published literature (preprint and peer-reviewed).

